# Estimating the true (population) infection rate for COVID-19: A Backcasting Approach with Monte Carlo Methods

**DOI:** 10.1101/2020.05.12.20098889

**Authors:** Steven J. Phipps, R. Quentin Grafton, Tom Kompas

## Abstract

Differences in COVID-19 testing and tracing across countries, as well as changes in testing within each country overtime, make it difficult to estimate the true (population) infection rate based on the confirmed number of cases obtained through RNA viral testing. We applied a backcasting approach, coupled with Monte Carlo methods, to estimate a distribution for the true (population) cumulative number of infections (infected and recovered) for 15 countries where reliable data are available. We find a positive relationship between the testing rate per 1,000 people and the implied true detection rate of COVID-19, and a negative relationship between the proportion who test positive and the implied true detection rate. Our estimates suggest that the true number of people infected across our sample of 15 developed countries is 18.2 (5-95% CI: 11.9-39.0) times greater than the reported number of cases. In individual countries, the true number of cases exceeds the reported figure by factors that range from 1.7 (5-95% CI: 1.1-3.6) for Australia to 35.6 (5-95% CI: 23.2-76.3) for Belgium.

## Introduction

COVID-19, caused by SARS-CoV-2, was declared a pandemic by the World Health Organisation on 11 March 2020, after first being identified in China in December 2019. As of early May 2020 there are more than 4 million reported cases globally, with the number of reported fatalities approaching 300,000.

Since January 2020, researchers have used the reported cases of COVID-19, caused by SARS-CoV-2, from tests for the presence of RNA material in nasal secretions or sputum in individuals, to estimate the rate of infection within a population. In many countries, an inadequate number of both testing kits and testing facilities, coupled with restrictions on who can be tested, has meant that the number of confirmed cases as a proportion of the total population underestimates the true (population) infection rate. Quantifying the true infection rate is an urgent health priority because, as late as the middle of March 2020, it was observed, and widely cited, that the data collected until that time were unreliable as a means to estimate the true infection rate^1^. Multiple lines of evidence also suggest that COVID-19 may be much more widespread within the population than is suggested by the outcomes of the limited direct testing conducted to date^2-12^.

A complementary approach to testing for RNA material of the virus is serological testing for antibodies for COVID-19. Sero-surveys provide an estimate of the number of people in the sample who have antibodies to the virus at a given point in time. To provide an estimate of the level and trend of the true infection rate, such testing needs to be repeated regularly and for an appropriately stratified random sample of the population. A challenge with sero-surveys is that the tests are subject to both false positives and false negatives and, in the case of serological testing for COVID-19, tests have been made available without the oversight required to ensure sufficient quality and accuracy^13^. Consequently, some tests are performing inadequately^14^. Another difficulty with serological tests is that, if the true rate of infection is relatively low (say 1% or less), then the number of false positives or false negatives may render sero-surveys unreliable as means to estimate the true infection rate. This can be the case even if the test has a high sensitivity (proportion of those tested who have had the virus and who return a positive result) and high specificity (proportion of those tested who have not been infected with the virus and who return a negative result).

A statistical approach to estimate the true number of infections is to backcast and to infer the true infection rate in the past, based on the current reported fatalities due to COVID-19. This approach has been used by Flaxman et al.^5^ to estimate the true infection rates for 11 European countries, and to model the rates of infection with and without physical distancing measures. We apply our own backcasting approach to estimate the true infection rate for 15 countries, without the need to employ an epidemiological model. By comparing our estimates with the reported number of confirmed cases, we derived an implied true detection rate by country. Using Monte Carlo methods to sample parameter uncertainty, we provided 5-95% confidence intervals for our estimates.

## Results

We used our backcasting approach to study the progression of the COVID-19 pandemic within 15 countries: the 11 European countries studied by Flaxman et al.^5^, as well as Australia, Canada, South Korea and the USA. The estimated cumulative numbers of true infections within each of the 15 countries are shown in Figure 1, while the implied detection rates are shown in Figure 2. A summary is also presented in Table 1.

**Figure 1.**
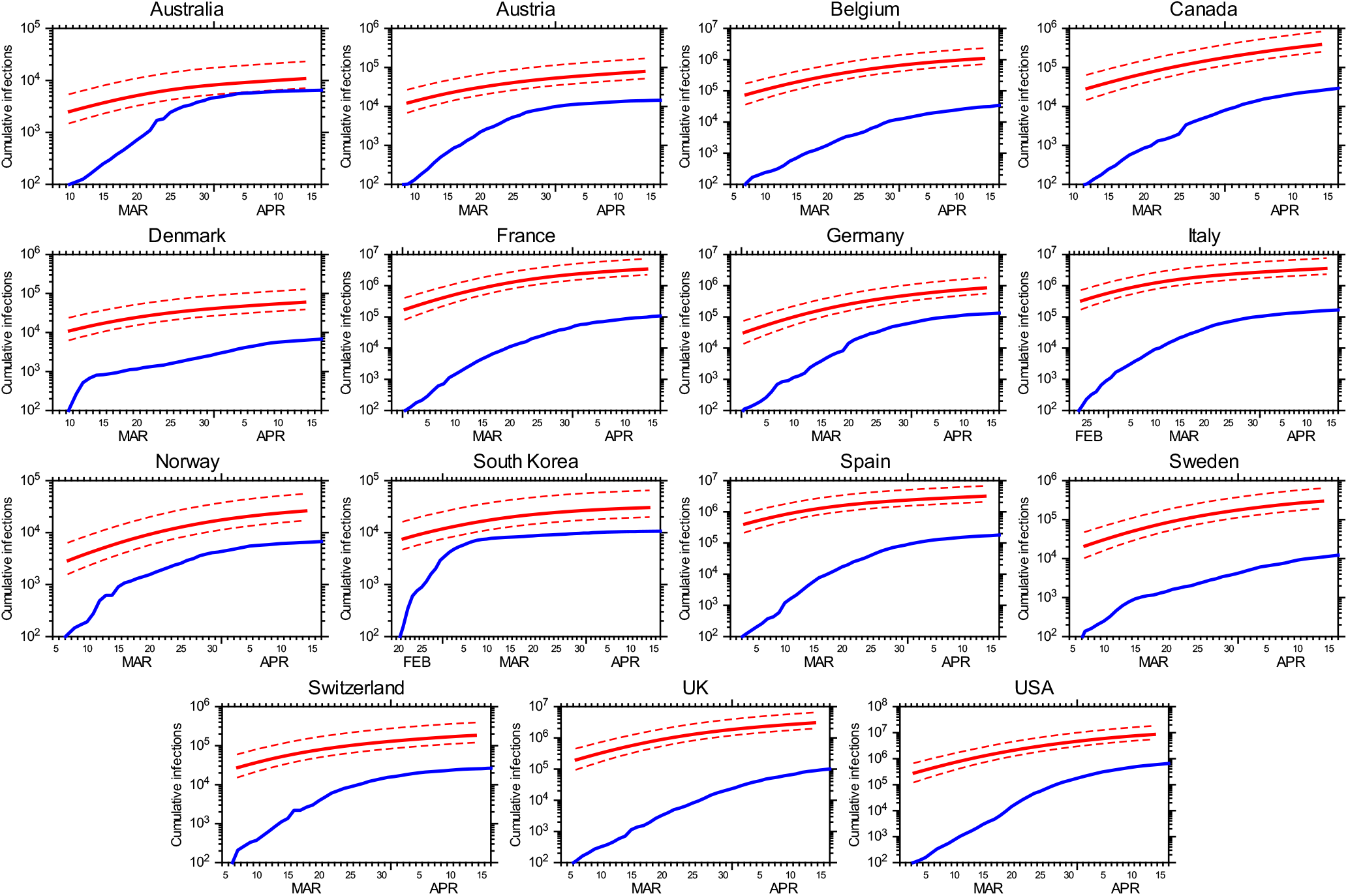
The cumulative number of COVID-19 infections for each country: the number of detected infections (blue) and the estimated true number of infections (red). For the true number, the median estimate and the 5-95% confidence interval are indicated by a solid line and dashed lines respectively. The data shown for each country begins on the day that the number of detected infections first reached or exceeded 100. Note that the vertical scale is different for each panel.

**Figure 2.**
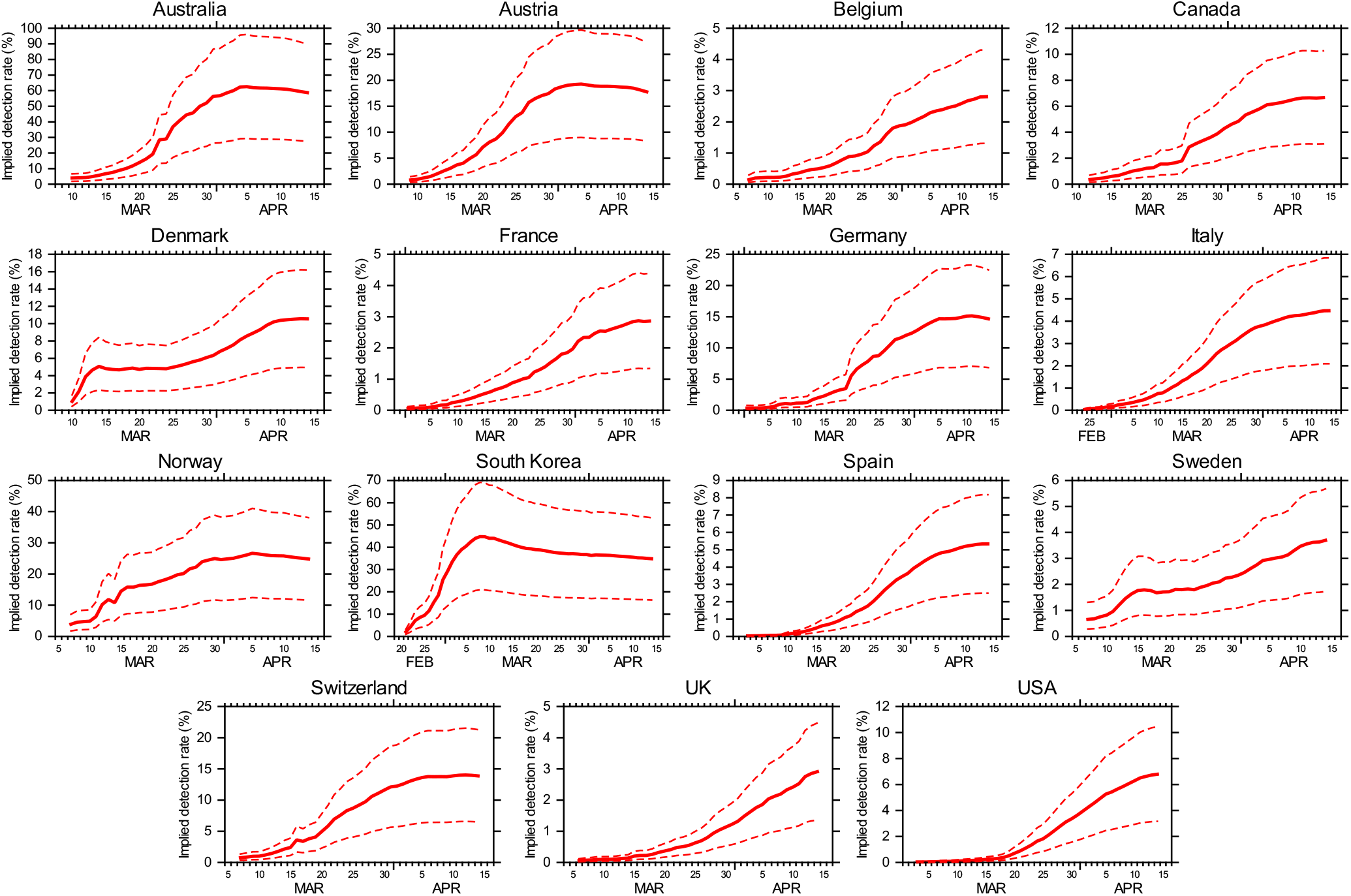
The implied detection rate for each country: the median estimate (solid line) and the 5–95% confidence interval (dashed lines). The data shown for each country begins on the day that the number of detected infections first reached or exceeded 100. Note that the vertical scale is different for each panel.

**Table 1.**
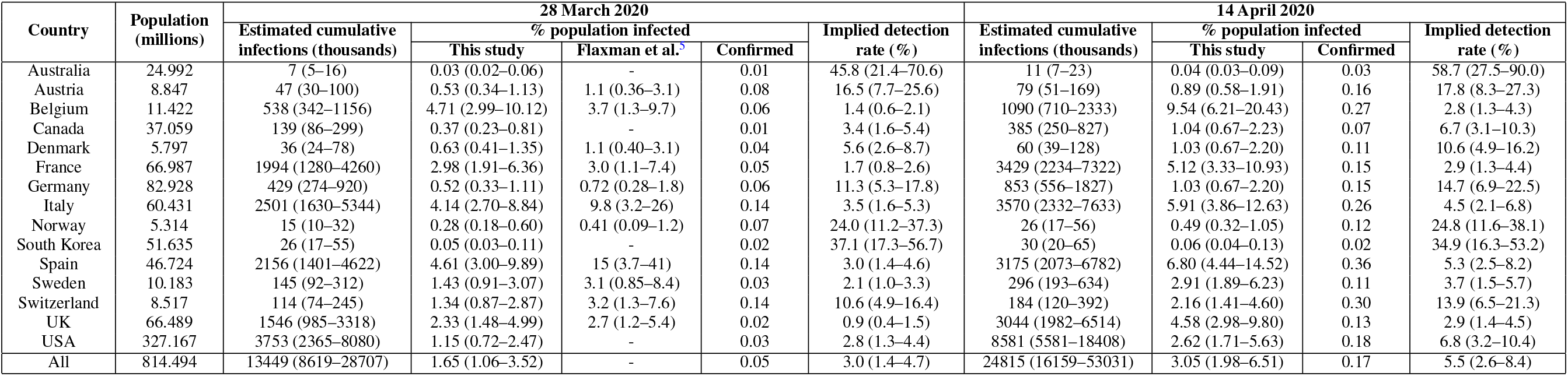
Statistics for each country as at 28 March 2020 and 14 April 2020: the estimated true cumulative number of infections (median and 5-95% confidence interval); the estimated cumulative percentage of the population to be infected (median and 5-95% confidence interval); the confirmed percentage of the population to have tested positive; and the implied detection rate (median and 5-95% confidence interval). For 28 March 2020, the estimated true cumulative number of infections according to Flaxman et al.^5^ (mean and 95% credible interval) is also provided for comparison. The data shown for “All” respresents aggregated statistics for all 15 countries. Population statistics are obtained from the European Centre for Disease Prevention and Control.

We find particularly low implied true detection rates (5-95% CI < 5%) for Belgium, France and the UK. Low implied true detection rates (5-95% CI < 10%) are also encountered in Sweden, Italy and Spain. By contrast, the implied true detection rates are high in countries that have low incidences of COVID-19 and/or have employed widespread testing, particularly South Korea and Australia.

We estimated the true cumulative number of infections as at 28 March 2020 and compared our estimates with those of Flaxman et al.^5^ for the same date (Table 1). For the 11 European countries for which a comparison is possible, our estimated infection rates tend to be slightly lower, particularly in the cases of Italy and Spain. Nonetheless, for every country, our 5-95% confidence intervals overlap with the 95% credible intervals of Flaxman et al.^5^. This demonstrates that our results are consistent with those generated using more complicated methods that involve the application of epidemiological models.

Our analysis covered 15 developed countries with a combined population of 814 million people. Between 28 March 2020 and 14 April 2020, we estimated that the total number of cumulative infections increased from 13.449 million (5-95% CI: 8.619-28.707 million) to 24.815 million (5-95% CI: 16.159-53.031 million). We, therefore, estimated that the fraction of the population to be infected increased from 1.65% (5-95% CI: 1.06-3.52%) to 3.05% (5-95% CI: 1.98-6.51%) over this period, with the implied true detection rate increasing from 3.0% (5-95% CI: 1.4-4.7%) to 5.5% (5-95% CI: 2.6-8.4%). These findings indicate that, on a global scale, COVID-19 is far more prevalent than is suggested by reported statistics, with the true number of infections exceeding the number of confirmed cases by a factor of 18.2 (5-95% CI: 11.9-39.0).

Our estimates of the implied true detection rate allowed us to explore the impact of testing on the rate of detection of COVID-19 infections in a population. A positive relationship is apparent between the implied true detection rate and the number of tests per 1,000 people (Figure 3), while a negative relationship is apparent between the implied true detection rate and the fraction of tests to return a positive result (Figure 4). The values of Spearman’s rank correlation coefficient are *ρ* = +0.52 (*p =* 0.048, *n =* 15) and *ρ* = −0.87 (*p =* 2.3 × 10^-5^, *n =* 15), respectively. Overall, the strength of these relationships demonstrates the benefits of sufficient testing to reliably estimate the level and trend of COVID-19 infection within a population.

**Figure 3.**
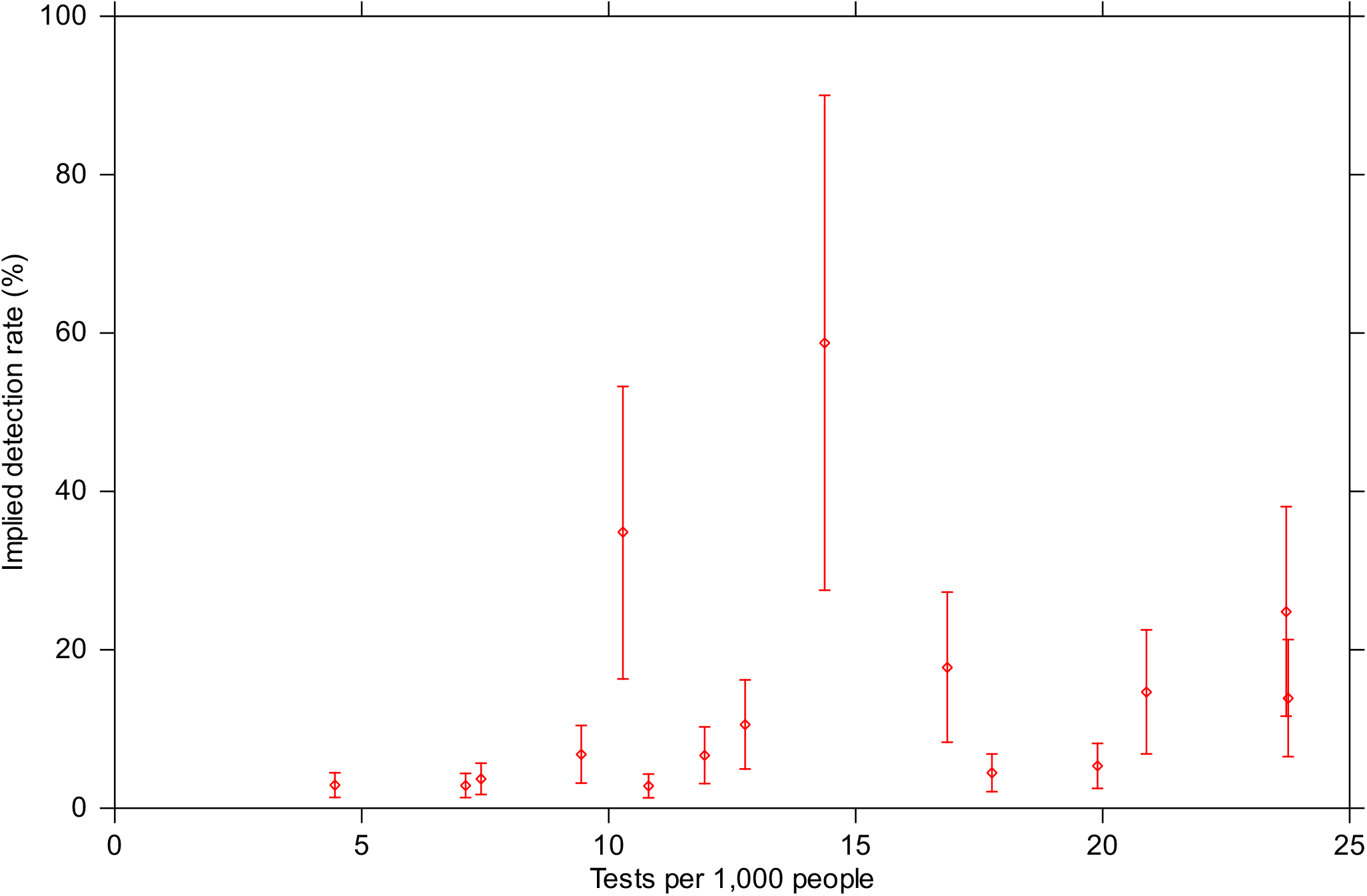
The implied true detection rate for each country versus the number of tests per 1,000 people: the median estimates (diamonds) and the 5-95% confidence intervals (vertical bars). All values are current as at 14 April 2020, except for Germany (testing data current as at 12 April 2020), Spain (testing data current as at 12 April 2020) and Sweden (testing data current as at 12 April 2020).

**Figure 4.**
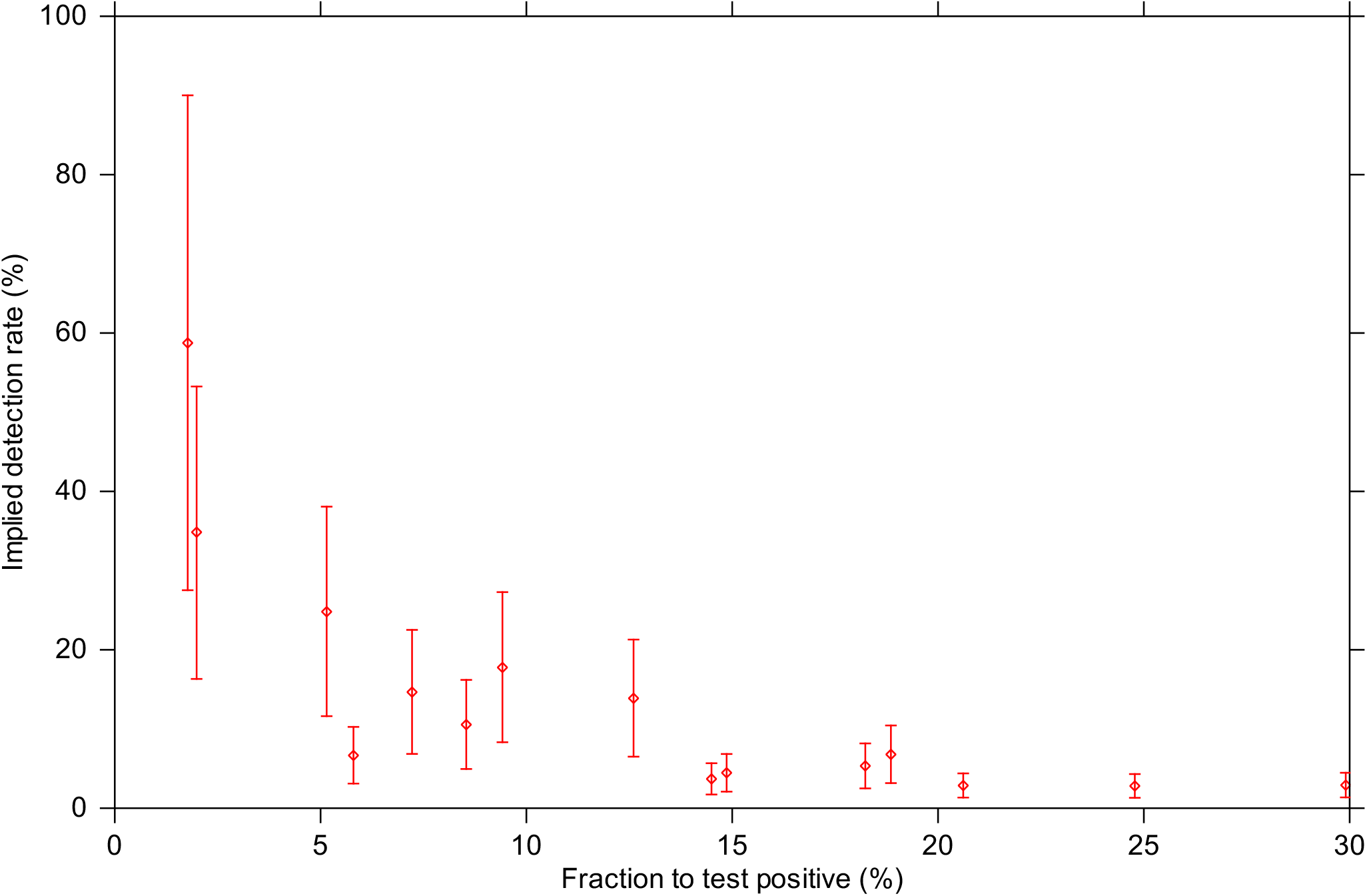
The implied true detection rate for each country versus the fraction of tests to return a positive result: the median estimates (diamonds) and the 5-95% confidence intervals (vertical bars). All values are current as at 14 April 2020, except for Germany (testing data current as at 12 April 2020), Spain (testing data current as at 12 April 2020) and Sweden (testing data current as at 12 April 2020).

## Discussion

Our backcasting approach is a valuable and easy-to-use method for estimating the true (population) infection rate wherever there is reliable data on the number of fatalities attributable to COVID-19. Our approach complements direct testing for the virus through nasal secretions or sputum. The difficulty is that direct testing for the virus in many countries is still not yet widespread, making inference of the true infection rate from the reported number of infections problematic.

Our approach complements attempts to estimate the true infection rate through sero-surveys coupled with stratified random sampling. The difficulty with sero-surveys is that some authorised tests perform poorly. Furthermore, even if serological tests have a high sensitivity and a high specificity, sero-surveys are unreliable if used to estimate the true infection rate when the infection rate is low, because the results will be confounded by the number of false positives and false negatives.

Backcasting has a number of advantages as a method when comparing estimated true infection rates across countries. Unlike direct testing for the virus, which varies greatly between countries because of the availability of testing kits and differences in testing protocols, backcasting only requires that fatalities as a result of COVID-19 be comparable. While it is true that countries do differ in how COVID-19 fatalities are recorded, we contend that these differences are much smaller than the variations in testing for the virus. Furthermore, the total number of COVID-19 fatalities can be estimated if recorded fatalities are only limited to hospital fatalities by, for example, comparing the overall fatality rate to a comparable period in previous years or including a proportion of the total number of fatalities occurring outside of hospitals from COVID-19-like symptoms.

The backcasting approach that we used to estimate the true infection rate is easily understood, transparent, scalable to a local, regional or a national level, and can be readily updated on a daily basis using data that has already been reported. Furthermore, it makes no assumptions with regard to how the number of COVID-19 infections has evolved over time. In our view, the ease of use of backcasting has particular advantages in countries with little testing or limited capacity to forecast rates of infection.

Complementary to backcasting are fit-for-purpose epidemiological models that are much better suited to predicting future hospitalisations and fatalities under different policy scenarios. We contend, however, that epidemiological models are not as suitable as backcasting for estimating the true infection rates because of their data requirements, modelling assumptions and because epidemiological model parameters may not be well calibrated at a local or regional scale.

We find using our backcasting approach that, in many countries, COVID-19 infections are far more prevalent within the population than is suggested by the reported positive tests of RNA viral material. Our results, therefore, complement the estimated infection rates derived using a range of other techniques, including direct testing of entire communities, sero-surveys and epidemiological modelling^2-12^. A key advantage of our approach is that it also allows for direct comparison of infection rates over time and across countries.

A valuable finding of this study is that there is a positive relationship between the rate of testing within the population and the implied true detection rate. We also find a negative relationship between the implied true detection rate and the proportion of positive viral tests for those tested for COVID-19. This demonstrates both the importance and the benefit of large-scale direct testing to determine the prevalence of COVID-19 within a population. Large-scale and sufficient testing — including the testing of those who are asymptomatic — is, therefore, of critical importance to inform policy decisions about how to resource, and how to manage, the impacts of COVID-19 on public health, society and the economy.

Our backcasting approach applied to the 15 countries in our sample implies that the true infection rate is 18.2 (5-95% CI: 11.9-39.0) times larger than the rate implied from the number of reported cases of COVID-19. Thus, collectively for all 15 countries, we estimated that a cumulative number of 24.815 million people (5-95% CI: 16.159-53.031 million) were infected with COVID-19 as at 14 April 2020, as compared to the reported total of 1.360 million. In some countries with very low implied true detection rates, such as Belgium, France, Italy, Sweden and the United Kingdom, our median estimate is that the reported number of cases, as at 14 April 2020, represented less than 5% of the true number of cases. Thus, a key implication of our work, especially as most countries in the world have undertaken fewer tests per 1,000 people and have a lower capacity to test than the 15 developed countries in our sample, is that the number of people who are infected with, or who have recovered from COVID-19, is many times greater than the reported number of cases from viral testing.

## Methods

### Backcasting

A backcasting method was used to estimate the true cumulative number of infections. Following Flaxman et al.^5^, the time from infection to death is assumed to follow a Gamma distribution. If the mean time from infection to death is *μ* and the standard deviation is s, then the distribution of times from infection to death is assumed to follow Gamma(*α, β*) with *α =* (*μ*/s)^2^ and *β =* (*s*^2^*/μ*).

The Gamma distribution can be used to project the number of new daily fatalities backwards in time from the time to death to the time of initial infection. Let *N_f_* (*t*) be the number of new fatalities to occur on day *t*. If *f* (*x*; *α, β*) is the probability density function for the Gamma distribution and IFR is the infection fatality rate, then the number of new infections estimated to have occurred on day *t′* and to have resulted in fatalities on day *t* is given by

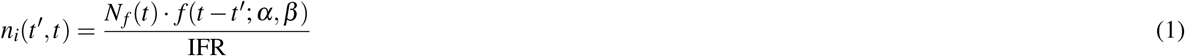

The estimated *total* number of new infections to have occurred on day *t′* is, therefore, given by summing the values of *n_i_*(*t*′, *t*) for all possible values of *t* > *t*′. This estimate is corrected because not all of the fatalities to arise from infections contracted on that day *t′* will have occurred yet. If *t*_0_ is the most recent day for which fatality statistics are available and *F* (*x*; *α, β*) is the cumulative distribution function for the Gamma distribution, then the estimated total number of new infections to have occurred on day *t′* is given by

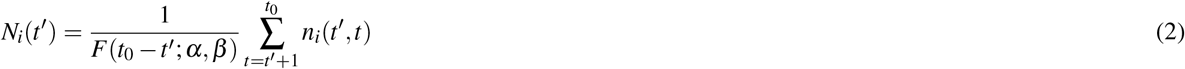

Equations 1 and 2 rely on the values of just three unknown parameters: (i) the mean time from infection to death; (ii) the standard deviation in the time from infection to death; and (iii) the infection fatality rate. In order to make the best use of published epidemiological data for COVID-19, we take the mean time from infection to death as being the sum of two other periods: the mean incubation period, and the mean time from development of symptoms to death. The standard deviations in each of these quantities can be estimated by following the approach of Flaxman et al.^5^ and taking the coefficients of variation as being 0.86 and 0.45, respectively. In practice, our backcasting method is based on the three parameters provided in Table 2.

Monte Carlo methods were used to sample parameter uncertainty, with the best estimate and uncertainty range for each parameter shown in Table 2. An ensemble with 10,000 members is generated with random draws from within the uncertainty range for each parameter using a triangular probability distribution. The uncertainty range of 0.2-1.6% for the infection fatality rate comes from Verity et al.^15^. The mean incubation period of 4.9 days, and the uncertainty range of 3.3-7.0 days, is derived by combining the estimates of Li et al.^8^ [5.2 (4.1-7.0) days] and Linton et al.^16^ [4.6 (3.3-5.8) days]. The mean time from symptoms to death of 15.8 days, and the uncertainty range of 11.8-19.2 days, is derived by combining the estimates of Linton et al.^16^ [13.8 (11.8-16.0) days] and Verity et al.^15^ [17.8 (16.9-19.2) days].

**Table 2.** The three parameters used in the backcasting exercise, including the best estimate and the uncertainty range.

| **Parameter** | **Units** | **Best estimate** | **Uncertainty range** |
| --- | --- | --- | --- |
| Infection fatality rate | % | 0.9 | 0.2–1.6 |
| Mean incubation period | days | 4.9 | 3.3–7.0 |
| Mean time from symptoms to death | days | 15.8 | 11.8–19.2 |

The implied true detection rate was calculated by dividing the cumulative reported number of infections for each country by our estimates of the true cumulative number of infections.

### Statistics

We used Spearman’s rank correlation coefficient to assess correlation in this study as it is a non-parametric measure that tests for a monotonic relationship, rather than a linear relationship, between two variables. The statistical significance of Spearman’s rank correlation coefficient *ρ* is tested by calculating the *t*-statistic using

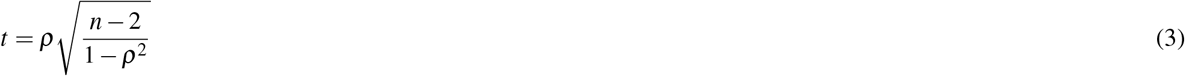

Under the null hypothesis of statistical independence, *t* can be assumed to be distributed as Student’s *t*-distribution with *n* − 2 degrees of freedom^17^. In this study, all the tests performed were two-tailed.

### Data

For each country, the population, the number of new infections reported each day, and the number of new fatalities reported each day are obtained from the European Centre for Disease Prevention and Control (https://www.ecdc.europa.eu/en/covid-19-pandemic). The cumulative number of tests performed per capita in each country is obtained from Our World in Data (https://ourworldindata.org/coronavirus).

We collected data for three other locations: New Zealand, Singapore and Taiwan. Gaps in the European Centre for Disease Prevention and Control data set prevented us from including New Zealand or Taiwan in our analysis. In the case of Singapore, our backcasting approach estimated an implied detection rate of 177.2% (5-95% CI: 82.9-271.2%) as at 14 April 2020. By definition, the detection rate cannot exceed 100%. While this upper limit lies within our 5-95% confidence interval, our results nonetheless suggest that there are unique characteristics to the nature of the COVID-19 pandemic in Singapore. We hypothesise that our analysis is affected by different mortality rates within its infected population; in particular, by the end of April 2020, ~90% of reported cases in Singapore were amongst its migrant worker population, who are young and healthy adults and who have, therefore, exhibited a very low mortality rate^18,19^. For this reason, we omitted Singapore from our analysis. Nonetheless, we note that including Singapore in our sample does not change our findings of a positive relationship between the implied detection rate and the number of tests per 1,000 people (*ρ* = +0.39, *p =* 0.14, *n* = 16) or of a negative relationship between the implied detection rate and the fraction of tests to return a positive result (*ρ* = −0.89, *p* = 5.1 × 10^−6^, *n* = 16).

### Data Availability

The datasets generated and analysed in this study are available at https://zenodo.org/record/3821525.

### Code Availability

The source code generated during this study is available at https://zenodo.org/record/3821525.

## Acknowledgements

We acknowledge the European Centre for Disease Prevention and Control and Our World in Data for making data on the COVID-19 pandemic freely available. We also acknowledge use of the PyFerret program to generate the graphics in this paper.

## Author contributions statement

R.Q.G. conceived of the approach. R.Q.G and S.J.P. conceived the experiments. S.J.P. conducted the experiments and analysed the results. R.Q.G and S.J.P interpreted the results. All authors wrote the manuscript.

## Correspondence

Correspondence and requests for materials should be addressed to S.J.P.

